# Secondary pneumonia in critically ill ventilated patients with COVID-19

**DOI:** 10.1101/2020.06.26.20139873

**Authors:** Mailis Maes, Ellen Higginson, Joana Pereira-Dias, Martin Curran, Surendra Parmar, Fahad Khokhar, Delphine Cuchet-Lourenço, Janine Lux, Sapna Sharma-Hajela, Benjamin Ravenhill, Razeen Mahroof, Amelia Solderholm, Sally Forrest, Sushmita Sridhar, Nicholas Brown, Stephen Baker, Vilas Navapurkar, Gordon Dougan, Josefin Bartholdson Scott, Andrew Conway Morris

**Affiliations:** Cambridge Institute of Therapeutic Immunology & Infectious Disease (CITIID), Department of Medicine,University of Cambridge, Cambridge, United Kingdom; Public Health England, Clinical Microbiology and Public Health Laboratory, Addenbrookes Hospital, Cambridge, United Kingdom; Department of Medicine, University of Cambridge, Cambridge, United Kingdom; John Farman ICU, Addenbrookes Hospital, Cambridge, United Kingdom; Wellcome Sanger Insitute, Hinxton, United Kingdom

**Author notes:** Corresponding author Andrew Conway Morris, Division of Anaesthesia, Department of Medicine, Level 4, Addenbrooke’s Hospital, Hills Road, Cambridge, CB2 0QQ. Contributed equally to the paper (Mailis Maes, Ellen Higginson, Joana Pereira-Dias). Author statement We studied the microbial composition and diagnostic microbiology results of bronchoalveolar lavage (BAL) samples taken from adult COVID-19 patients and COVID-19 negative patients receiving mechanical ventilation in 3 hospital ICUs. We observed that although COVID-19 patients had a greater incidence of VAP, the associated causative pathogens were similar in both groups.

**Keywords:** COVID-19, SARS-CoV-2, coinfections, microarray, VAP, ICU

## Abstract

**Background:** Pandemic COVID-19 caused by the coronavirus SARS-CoV-2 has a high incidence of patients with severe acute respiratory syndrome (SARS). Many of these patients require admission to an intensive care unit (ICU) for invasive artificial ventilation and are at significant risk of developing a secondary, ventilator-associated pneumonia (VAP).

**Objectives:** To study the incidence of VAP, as well as differences in secondary infections, and bacterial lung microbiome composition of ventilated COVID-19 and non-COVID-19 patients.

**Methods:** In this prospective observational study, we compared the incidence of VAP and secondary infections using a combination of a TaqMan multi-pathogen array and microbial culture. In addition, we determined the lung microbime composition using 16S RNA analyisis. The study involved eighteen COVID-19 and seven non-COVID-19 patients receiving invasive ventilation in three ICUs located in a single University teaching hospital between April 13^th^ 2020 and May 7^th^ 2020.

**Results:** We observed a higher percentage of confirmed VAP in COVID-19 patients. However, there was no statistical difference in the detected organisms or pulmonary microbiome when compared to non-COVID-19 patients.

**Conclusion:** COVID-19 makes people more susceptible to developing VAP, partly but not entirely due to the increased duration of ventilation. The pulmonary dysbiosis caused by COVID-19, and the array of secondary infections observed are similar to that seen in critically ill patients ventilated for other reasons.

## Background

Pandemic COVID-19 is associated with a high number of patients suffering from severe acute respiratory syndrome (SARS). Such patients can spend significant periods of time in intensive care units (ICU), with >80% of patients admitted to ICU requiring invasive mechanical ventilation [1,2]. Critically ill patients are at high risk of nosocomial pneumonia, especially when ventilated [3]. The reasons for this includes breach of natural defences by invasive devices [4], sedation and impairment of coughing and mucociliary clearance, and the immunoparetic effects of critical illness [5,6]. Early reports indicated that critically ill patients infected with SARS-CoV-2 had a high prevalence of nosocomial pneumonia, especially ventilator-associated pneumonia (VAP) [7]. This is likely influenced by the widespread use of corticosteroids and empiric immunosuppressive medication together with increased prevalence of co-morbid conditions [7] and the prolonged duration of artificial ventilation [2].

The management of critically ill patients with COVID-19 requires the identification, or exclusion, of bacterial, viral or fungal pathogens which may be present as co-infections on presentation or arise later in the course of the disease. Ventilator-associated pneumonia (VAP) can be challenging to diagnose as a range of non-infectious diseases may mimic the clinical picture of radiographic infiltrates, systemic inflammation and impaired oxygenation that typifies VAP [8]. To limit overdiagnosis and facilitate appropriate antimicrobial therapy in VAP, most centres use culture-based approaches [9,10]. However, molecular tests to detect multiple pathogens (viruses and bacteria) are becoming more accessible and may further reduce unnecessary antimicrobial therapy [11]. Additionally, the choice of diagnostic sample is critical and directed bronchoscopy can limit contamination from the proximal airway [12]. An observation that the rate of VAP amongst patients with COVID-19 appeared to be higher than our background rate led to the institution of a minimally-aerosol generating bronchoscopic sampling procedure to seek to minimise over-diagnosis inherent in endo-tracheal aspirate-based sampling [12].

In this study, we aimed to identify and compare the distribution of secondary infections and VAP in critically ill ventilated COVID-19 patients against ventilated non-SARS-CoV-2 infected patients. We performed conventional microbiology, multi-pathogen molecular testing using a TaqMan array card developed and validated previously [13], and assessed the composition of the bacterial lung microbiome in bronchoalveolar lavage (BAL) samples of COVID-19 positive and COVID-19 negative patients in the same hospital over a certain time period.

## Materials and methods

### Setting and study design

This study was a performed in three adult ICUs in Addenbrooke’s Hospital, Cambridge, UK, consisting of a liver/general unit, neurotrauma unit and a surge capacity COVID unit. Units had routinely audited ventilator bundles in place, which included sub-glottic suction tubes, mandated twice daily oral hygiene with fluoride toothpaste, daily sedation holds and head of bed elevation. Patients from April 13^th^ to May 7^th^ were included in the study if the treating clinician was undertaking BAL for the diagnosis of respiratory infection in a ventilated patient. All patients had X-ray infiltrates and features of systemic inflammation (raised white cell count, temperature >38°C, raised C-reactive protein and/or raised serum pro-calcitonin levels).

BALs were conducted in accordance with a modified unit protocol designed to minimise aerosol generation. Briefly, staff members wearing appropriate personal protective equipment (PPE) inserted a pre-loaded bronchoscope through an endotracheal tube catheter mount. The bronchoscope was wedged in a sub-segmental bronchus corresponding to the region of maximal infiltrate on plain radiography. Up to 200ml of saline was instilled and aspirated.

### Diagnostics

Samples for routine microbiology were processed according to the UK Standards for Microbiology Investigations [14]. Any significant growth with a CFU of >10^4^/mL was identified by MALDI-ToF mass spectrometry.

### RNA/DNA extraction and SARS-CoV-2 qPCR

500µl of BAL was subjected to RNA/DNA extraction following an existing method (14). Viscous samples were first treated with 10% v/v mucolysin, before 500µl lysis buffer (25mM Tris-HCL+ 4M Guanidine thiocyanate with 0.5% b-mercaptoethanol) and glass beads were added to each sample. Tubes were vortexed, and 100% analytical grade ethanol was added to a final concentration of 50%. After a 10 min incubation, 860µl of lysis buffer (containing MS2 as an internal extraction and amplification control) was added. This was then run over an RNA spin column as previously described [15]. SARS-CoV-2 specific real-time RT-PCR was performed and interpreted as previously described [15].

### TaqMan multi-pathogen array

Custom designed TaqMan Array Cards (TAC; Thermo Fisher Scientific) targeting 52 different common respiratory pathogens, were used to test for secondary infections as previously described [13]. Fifty microlitres of extracted nucleic acid was used in a 200 μl final reaction volume with TaqMan™ Fast Virus 1-step Master Mix (Thermo Fisher Scientific), and cards were run on the QuantStudio 7 Flex platform (Thermo Fisher Scientific) as per the manufacturer’s instructions. Detection of a clear exponential amplification curve with a Cycles to Threshold (CT) value ≤ 32 for any single gene target was reported as a positive result for the relevant pathogen. In those patients who had BALs obtained more than once, new pathogens identified in later samples were added to the results of the initial array.

### VAP diagnosis

The definition of VAP was adapted from the European Centre for Disease Control criteria [16]. VAP was determined to be present in those patients with clinical evidence of pulmonary inflammation, radiographic evidence of lung infiltrates and detection of significant amounts of pathogenic bacterial or fungal species. Clinically significant amounts of pathogen were defined as those detected at a CT value ≤ 32 and/ or microbial growth on culture of ≥10^4^ CFU/ml. Low lung pathogenicity organisms (*Enterococcus spp*., *Candida albicans*, non-pneumococcal *Streptococci* and coagulase negative *Staphylococci*) were reported but not considered a component of VAP [17]. *Herpesviridae* (Herpes simplex, cytomegalovirus and Epstein-Barr virus) were reported but were considered to be reactivations and not considered a component of VAP [18].

### 16S Nanopore sequencing

Extracted nucleic acids were concentrated using AMPure XP beads (Beckman Coulter) and 16S DNA libraries prepared using the 16S Barcoding Kit SQK-16S024 (Oxford Nanopore Technologies) as per the manufacturer’s instructions.Final DNA libraries were loaded onto FLO-MIN106D R9.4.1 flow cells and sequencing was performed on a GridION Mk1 for ∼36 hours with high accuracy basecalling enabled. The resulting fastq files were de-multiplexed with guppy_barcoder v3.6.0 using the -- require_barcodes_both_ends and --trim_barcodes flags. Porechop v0.2.4 (https://github.com/rrwick/Porechop) was used to trim adapter and barcode sequences and Nanofilt v2.6.0 (De Coster, et. al., 2018) was used to filter the reads by length, 1,400 – 1,600 bps and a quality score of 10. Reads were classified against the Silva 132 99% OTUs 16S database using Kraken2 [19]. Microbial diversity analyses were carried out in R using packages vegan [20] and metacoder [21].

## Results

During the COVID-19 outbreak we have used a combination of TaqMan multi-pathogen array and conventional microbial culture to investigate secondary infections associated with patients undergoing ventilation. In our ICU, VAP was suspected in 82% of ventilated patients with COVID-19 and confirmed by culture or TAC in 49% of these patients, giving an incident density of 26 per 1000 ventilator days for confirmed VAP and 52 per 1000 ventilator days for suspected VAP. We report in more detail the diagnoses made from protected lower respiratory samples analysed by these combined culture-based and molecular techniques.

34 BAL samples were taken from 25 patients, of which the clinical and demographic data are summarized in Table 1; five patients were sampled on two occasions and two patients sampled on three occasions over the study period. Nineteen of these samples were SARS-CoV-2 positive by RT-PCR, with a mean CT of 28.3. Fifteen samples tested negative for SARS-CoV-2; however, six of these samples came from patients previously diagnosed with COVID-19 by RT-PCR (Table 2).

**Table 1.** Clinical and demographic features of reported population.

| Characteristic |  |
| --- | --- |
| Median age (range) | 57 (31-73) |
| Female n(%) | 9 (36%) |
| Antibiotics at time of bronchoscopy n(%) | 24 (96%) |
| Median FiO2 prior to bronchoscopy (range) | 0.5 (0.25-0.9) |
| Alive at end of study<br>(remain in ICU) | 17 (68%)<br>11 |
| Median Duration of ventilation: days (range) | 17 (2-42) |
| Median duration of ICU admission: days<br>(range) | 19 (2-44) |
| Immunocompromised* n(%) | 6 (24%) |

\*Immunocompromised patients were defined as having active haematological malignancy, neutropaenic malignancy, solid organ or bone marrow transplant and receipt of immunosuppressive medication including corticosteroids for >1 week prior to hospital admission.

**Table 2.**
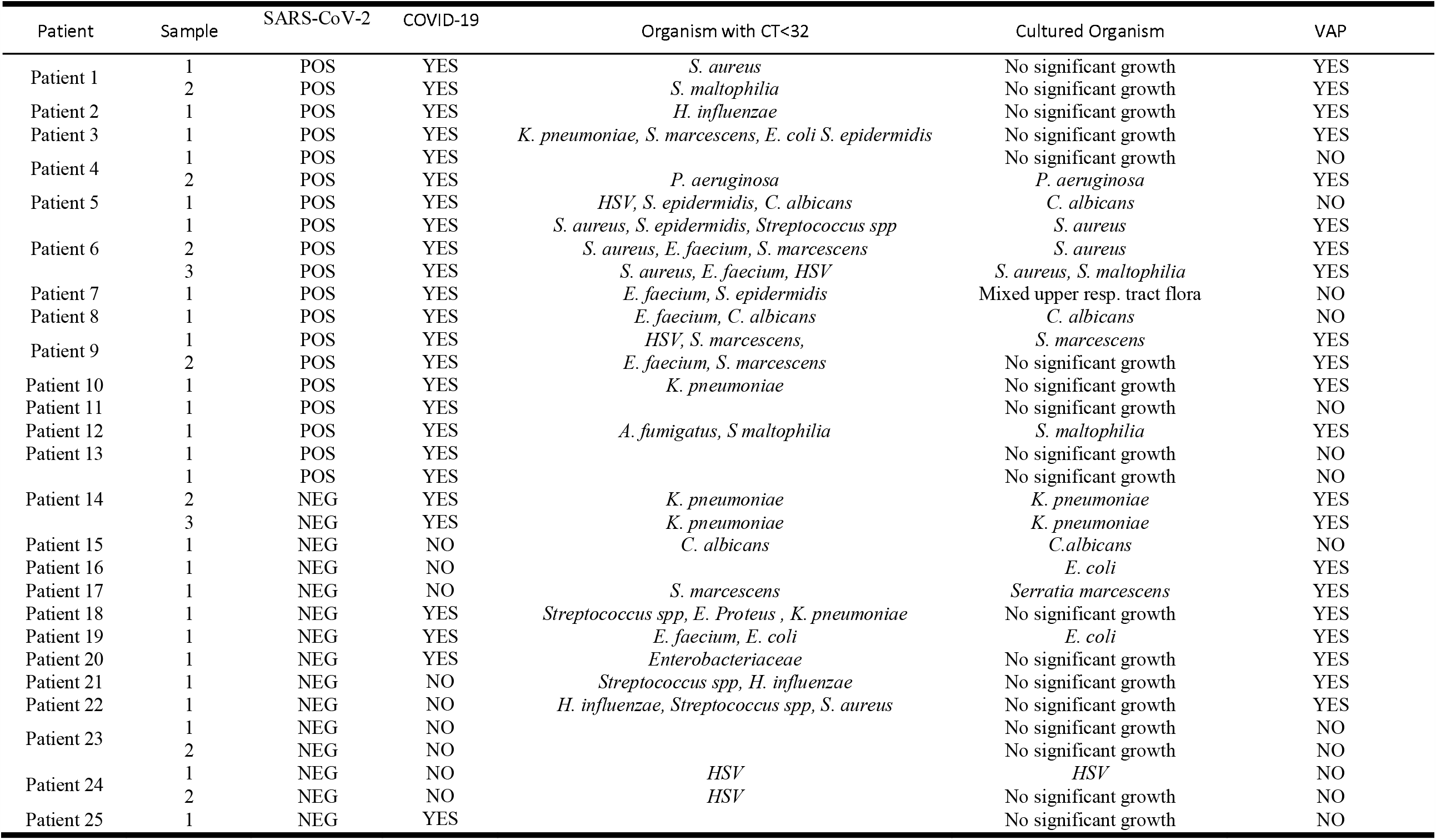

Amongst the 14 patients with a SARS-CoV-2 positive RT-PCR at time of sampling, nine were confirmed to have VAP on the basis of clinical features and detection of a lung pathogen at a CT ≤ 32 or by microbial culture >10^4^ CFU/ml. Of these, one patient incurred two episodes of VAP, first with *S. aureus* and later with *S. maltophilia*. Of the seven patients without COVID-19, four developed confirmed VAP. Three of the four patients previously diagnosed with COVID-19 but with negative SARS-CoV-2 RT-PCR at the time of sampling also were confirmed as VAP. (The co-infecting organisms are shown in Table 2).

Notably, there was no significant difference in the concentration (CT) or distribution of organisms between the COVID-19 positive and negative patients. Although not classed as VAP here, a number of organisms (*Enterococcus spp*., *Candida albicans*, non-pneumococcal *Streptococci*) and reactivated viruses (Herpes simplex), normally regarded as being of low pathogenic potential, were detected in patients (Table 2). In some cases, these were detected alongside classical lung pathogens, whereas in other patients they were the sole organisms detected. There was no clear difference in the prevalence of low-pathogenicity organisms between patients positive or negative for SARS-CoV-2 The mean number of organisms detected by TAC in the COVID positive patients was 1.8 organisms/patient (range 0 − 5), whereas the equivalent number in the COVID negative group was 1.1 organisms/patient (range 0 − 3).

To investigate changes in the lung microbiota in the COVID-19 positive and negative patients we performed 16S rRNA sequencing on a subset of BAL samples. In general, bacteria detected by TAC or conventional microbiology were abundantly identified in samples by 16S sequencing (Figure 1). When comparing COVID-19 positive to COVID-19 negative patients, there was no specific taxon that was more prevalent in either group. Additionally, the microbiomes of COVID-19 positive patients were not significantly different in either the species richness (alpha diversity) or the microbial composition (beta diversity) to those of COVID-19 negative patients.

**Figure 1.**
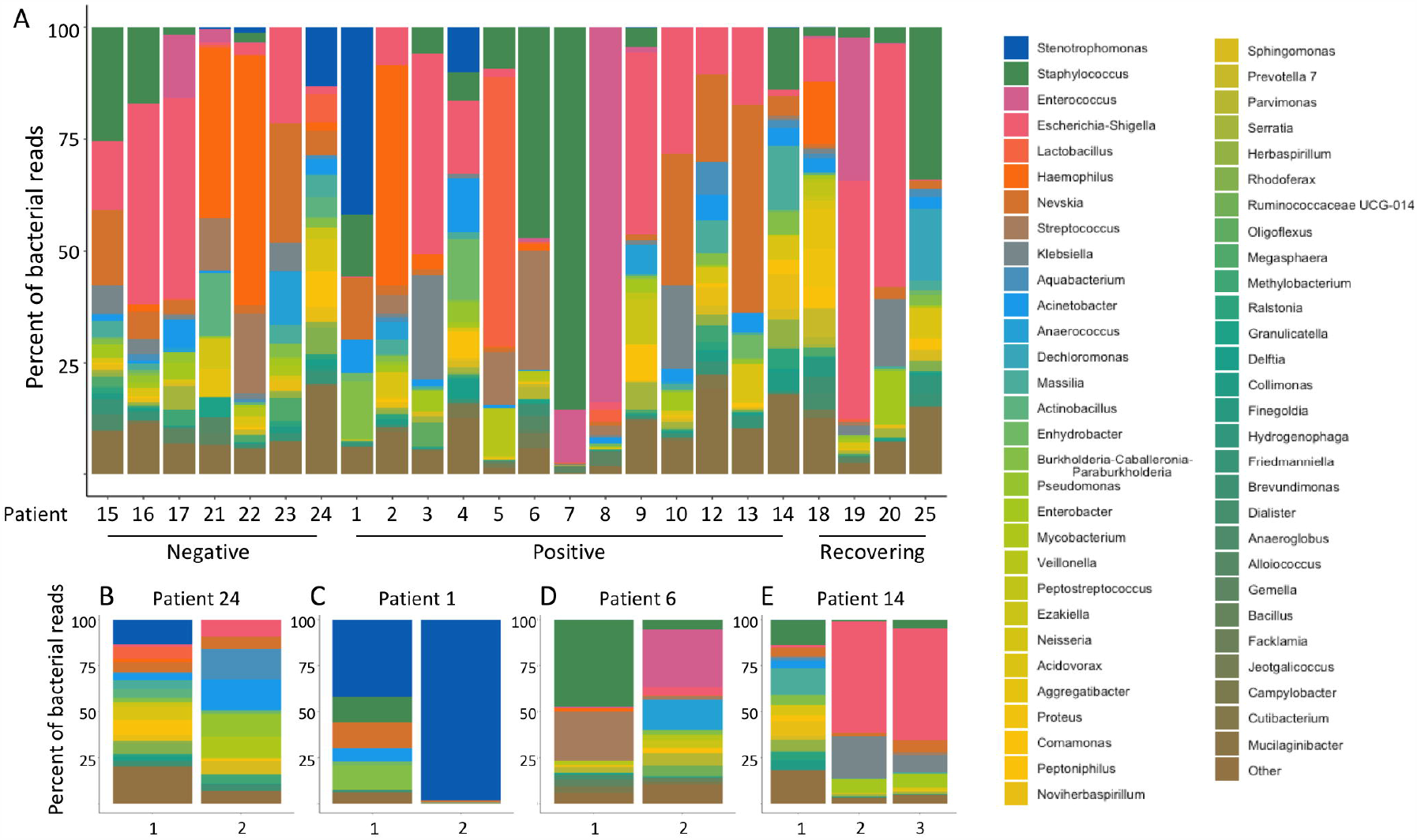
Microbial composition of BAL samples from SARS-CoV-2 positive and negative patients. Bacterial 16S genes were sequenced and classified to the genus level using Kraken2. The percent of reads mapping to each genus is shown for individual samples from each patient (A), and longitudinal samples (1, 2 or 3) from individual patients (B, C, D, E). Individuals were classified as either COVID-19 negative, COVID-19 positive, or recovering (previously diagnosed with COVID-19 but SARS-CoV-2 negative at time of sample).

To investigate changes in the microbiota over the course of infection, we next looked at the microbial composition of BAL samples in individual patients over time. Two patients diagnosed with VAP (patients 1 and 24) showed decreasing species richness over time, as the bacterial pathogen implicated in the illness became the predominant microbe present. For patient 6, the microbial composition shifted significantly over time, as *Enterococcus* took over from *Staphylococcus* as the most predominant pathogen. The microbiome composition of patient 24, who was both VAP and COVID-19 negative, was largely stable over time. In general, the lung microbiomes of patients who did not have VAP at the time of sampling (sample 1 from patient 14 and both samples from patient 24) were more diverse than samples from patients who had been diagnosed with VAP.

## Discussion

COVID-19 is a very new disease in the human population and this has led to an increase in the number of patients in need of active sustained ventilation, which in turn introduces an increased risk of VAP. COVID-19 can present in many different severe manifestations and reports of co-infections vary [22,7]. However, often these reports suffer from a lack of clarity around the severity of illness, location of patients (critical care vs non-critical care), timing of sampling relative to onset of disease and, where applicable, the use of mechanical ventilation [22]. Here, we report on the most severely affected COVID-19 patients who required clinical management on an ICU with mechanical ventilation. We found a high incident density of confirmed (26/1000 ventilator days) and suspected (52/1000 ventilator days) VAP in COVID-19 patients. This is greater than the previously reported rates from units with similar admission profiles and use of ventilator bundles, where incident densities were 6-14/1000 ventilator days for confirmed and 12-32/1000 ventilator days for suspected VAP [23]. Although incident density can correct for duration of ventilation to some extent, it is imperfect and long-staying patients may display different features from shorter staying patients [24]. However, even when compared to reports of patients staying for >14 days [23], incident density for COVID-19 patients remains high.

At the lung microbiome level, we observed no difference in the composition of organisms between COVID-19 positive and COVID-19 patients who developed VAP. Similarly, the causative pathogens identified using molecular and culture methods, were comparable in both the COVID-19 positive and negative patients. Reassuringly, antibiotic susceptibility of the causative pathogens was similar in the two groups (data not shown) and this meant that conventional antimicrobial regimens could be used. We did detect by TAC assay *Aspergillus fumigatus* in one patients, with supportive clinical and radiographic features, suggesting genuine fungal infection. This is notable as there is increasing recognition of fungal infections amongst patients with viral pneumonitides [25]. Our data support the concept that COVID-19, in common with other critical illness syndromes requiring mechanical ventilation, induces a pulmonary dysbiosis, leading to overgrowth of enteric and respiratory organisms, many of which are of low pathogenic potential[26,27]. These observations likely reflect intercurrent antimicrobial therapy and a degree of immunoparesis which is also observed in other critical illness states. Systemic inflammation, including activation of complement and release of C5a is a hallmark of severe COVID-19 [7, 28], with excessive C5a release being a key driver of innate immune dysfunction in critically ill patients and predictor of subsequent infections [23,5].

Studies reporting on VAP in COVID-19 patients are limited and have not reported the microbiological or diagnostic details of the case [7]; however, it has been suggested that bacterial pneumonia may be facilitated by the use of corticosteroids and empiric immunosuppressive medication. In our setting these medications are not commonly used, yet there remains a high prevalence of bacterial VAP in COVID-19 patients. Although VAP in COVID-19 may present problems of quantity, we did not find evidence in this report of a qualitative difference. Indeed, the microbial profiles of ventilated patients with active SARS-CoV-2, those who had cleared SARS-CoV-2 and those who never had the viral infection were similar on both targeted TAC and 16S rRNA sequencing. Our patients demonstrated similar profiles to those reported by other groups investigating the pulmonary microbiome of ventilated patients [26,29]. The factors which lead to pulmonary dysbiosis in critical illness remain incompletely understood, but may include intercurrent antibiotic use, enteric translocation, pulmonary immune dysfunction and altered clearance [30, 27]. We acknowledge the sample size limitations with our observations and suggest larger studies from distinct geographic locations may help fully understand the risk of developing secondary bacterial infections in patients with severe COVID-19.

## Conclusion

COVID-19 makes people more susceptible to developing VAP, partly but not entirely due to the increased duration of ventilation. The change in lung microbiome and causes of secondary infection are similar to those seen in critically ill patients ventilated for other reasons. Careful sampling of the respiratory tract whilst minimising contamination from the proximal tract, in combination with sensitive diagnostic testing to reduce the risk of false negative cultures will aid antimicrobial optimisation in patients with COVID-19.

## Ethical approval and consent to participate

The *TaqMan multi-pathogen array* has been adopted as a routine clinical service in our institution following a previous evaluation study [13]. The use of discard samples surplus to that required for clinical testing, and anonymised data review were conducted under the consent waiver granted by Leeds West NHS Research Ethics Committee (ref: 20/YH/0152).

## Data Availability

Raw sequencing data is available in SRA under project number PRJNA642012

https://www.ncbi.nlm.nih.gov/bioproject/PRJNA642012

## Acknowledgments

Our thanks go out to all the ICU clinicians, nurses and physiotherapists who have ensured proper patient care and sample management even in these hard times, as well as to PHE microbiologists for running standard microbiology tests. Furthermore, the authors wish to thank Mark Wills for his help as Biological Safety Officer to ensure we work in a safe environment. We also thank Estée Török and Ian Goodfellow for access to consumables and the GridION for Nanopore sequencing, and Satpal Ubhi for help with clinical data collection. Finally, we wish to acknowledge our funders the NIHR BRC. In addition we would like to acknowledge the grant awarded by Addenbrooke’s Charitable Trust for the initial study of the clinical utility of the TaqMan microarray [13].

## Authors contributions

MM-sample processing, data analysis and manuscript writing; EEH-sample processing, data analysis and manuscript writing; JPD-sample processing and manuscript editing; DCL,JL,AS, SF,SS -sample processing; FK-sample processing and data analysis; SSH, BR, RM-clincial data collection MC,SP, NB-diagnostic data analysis; SB manuscript editing; VN conceived study and sample acquisition: GD-conceived study and manuscript editing; JBS conceived study, sample processing and manuscript editing; ACM-conceived study, manuscript writing, sample acquisition, data analysis and clinical data collection.

## Consent for publication

All authors read and approved the final version for publication.

## Availability of supporting data

Raw sequencing data is available upon request.

